# Inference of SARS-CoV-2 spike-binding neutralizing antibody titers in sera from hospitalized COVID-19 patients by using commercial enzyme and chemiluminescent immunoassays

**DOI:** 10.1101/2020.09.07.20188151

**Authors:** Arantxa Valdivia, Ignacio Torres, Víctor Latorre, Clara Francés-Gómez, Eliseo Albert, Roberto Gozalbo-Rovira, María Jesús Alcaraz, Javier Buesa, Jesús Rodríguez-Díaz, Ron Geller, David Navarro

## Abstract

**Background:** Whether antibody levels measured by commercially-available enzyme or chemiluminescent immunoassays targeting the SARS-CoV-2 spike (S) protein can act as a proxy for serum neutralizing activity remains to be established for many of these assays.

**Objectives:** To evaluate the degree of correlation between neutralizing antibodies (NtAb) binding the SARS-CoV-2 Spike (S) protein and SARS-CoV-2-S-IgG levels measured by four commercial immunoassays in sera drawn from hospitalized COVID-19 patients.

**Patients and Methods:** Ninety sera from 51 hospitalized COVID-19 patients were assayed by a pseudotyped virus neutralization assay, the LIAISON SARS-CoV-2 S1/S2 IgG, the Euroimmun SARS-CoV-2 IgG ELISA, the MAGLUMI 2019-nCoV IgG and the COVID-19 ELISA IgG assays.

**Results:** Overall, the results obtained with the COVID-19 ELISA IgG test showed the highest agreement with the NtAb assay (*κ*, 0.85; 95% CI, 0.63-1). The most sensitive tests were the pseudotyped virus NtAb assay and the COVID-19 ELISA IgG assay (92.2% for both). Overall, the degree correlation between antibody titers resulting in 50% virus neutralization (NtAb_50_) in the pseudotyped virus assay and SARS-CoV-2 IgG levels was strong for the Euroimmun SARS-CoV-2 IgG ELISA (Rho = 0.73) and moderate for the remaining assays (Rho = 0.48 to 0.59). The kinetic profile of serum NtAb_50_ titers could not be reliably predicted by any of the SARS-CoV-2 IgG immunoassays.

**Conclusions:** The suitability of SARS-CoV-2-S-IgG commercial immunoassays for inferring neutralizing activity of sera from hospitalized COVID-19 patients varies widely across tests and is influenced by the time of sera collection after the onset of symptoms.

## 1. Background

Severe acute respiratory syndrome coronavirus 2 (SARS-CoV-2), a prototypical *Sarbecovirus*, causes Coronavirus disease 2019 (COVID-19) and is associated with significant morbidity and mortality [1,2]. SARS-CoV-2 neutralizing antibodies (NtAb) presumably play a pivotal role in preventing infection and may promote virus clearance [3,4]. In support of these assumptions, passive transfer of two mAbs blocking SARS-CoV-2 interaction with angiotensin-converting enzyme II receptor as monotherapy protected rhesus macaques from infection [5]. Moreover, transfusion of plasma from immune individuals with high NtAb titers seemed to be associated with improved clinical outcomes in critically ill COVID-19 patients [6,7]. Virus neutralization assays, either using live native SARS-CoV-2 virus, engineered SARS-CoV-2 pseudotyped viruses or replication-competent SARS-CoV-2 chimeric viruses [8-10] are cumbersome, require specialized facilities, and are time consuming. A large number of enzyme-linked (ELISA) or chemiluminescent immunoassays (CLIA) detecting antibodies that bind SARS-CoV-2 structural proteins have been commercialized [11]. Whether antibody levels measured by these serological assays can be used as a proxy for serum neutralizing activity is a relevant issue, which has been dealt with in several peer-reviewed or preprint studies [12-20], but demands further investigations.

## 2. Objectives

We focused on evaluating the degree of correlation between NtAb binding the SARS-CoV-2 S protein, which is known to elicit the most potent antibodies for virus neutralization [21,22], and SARS-CoV-2 IgG levels measured by four commercially-available semiquantitative immunoassays targeting the S protein in sera drawn from hospitalized COVID-19 patients.

## 3. Study design

### 3.1. Serum specimens and patients

A total of 90 sera from 51 non-consecutive patients with laboratory-confirmed SARS-CoV-2 infection by RT-PCR [23,24] that were admitted to Hospital Clínico Universitario of Valencia between March 5 to April 30, 2020, were included in the current study. The Demographic, clinical, and laboratory data of these patients are displayed in Table 1. Forty-one sera were obtained within the first two weeks (< day 15) after the onset of symptoms, at a median of 11 days (range, 5-14 days), and 49 afterward (≥15 days, at a median of 23 days; range, 15-41 days). Sequential specimens were available from 20 out of the 51 patients (median 3 specimens per patient; range 2 to 6). This study was approved by the Research Ethics Committee of Hospital Clínico Universitario INCLIVA (March 2020).

**Table 1.** Demographic, clinical and laboratory characteristics of patients with COVID-19

| **Parameter** | **Patients** |
| --- | --- |
| **Sex: Male/Female; no. (%)** | 32 (63)/ 19 (37) |
| **Age; median (range)** | 53 (21-77) |
| **Days of hospitalization; median (range)** | 17 (2-67) |
| **Co-morbidities; no. (%)** | 35 (69) |
| **Number of comorbidities; median (range)** | 1 (0-5) |
| Arterial hypertension | 23 (45) |
| Chronic renal disease | 2 (4) |
| Diabetes mellitus | 12 (24) |
| Dyslipidemia | 16 (31) |
| Ischemic cardiovascular disease | 4 (8) |
| Myocardial infarction | 2 (4) |
| Pulmonar disease<sup>a</sup> | 7 (14) |
| Tumor | 3 (6) |
| **Laboratory findings; median (range)<sup>a</sup>** |  |
| CRP (in mg/l) | 44 (0.8-273) |
| Ferritin (ng/ml) | 674 (2.5-2986) |
| Dimer-D (ng/ml) | 903 (91-5445) |
| Lactose dehydrogenase (U/l) | 666 (357-1328) |
| Interleukin-6 (pg/ml) | 1012 (4.6-5000) |
| Total lymphocyte count (*10<sup>9</sup>/L) | 1.15 (0.17-3.98) |
| <sup>a</sup>measurements in sera used for serological analyses reported in the current study. |  |

### 3.2. SARS-CoV-2 Neutralizing antibody assay

A Green fluorescent protein (GFP) reporter-based pseudotyped virus neutralization assay using a non-replicative vesicular stomatitis virus (VSV lacking the G protein) backbone coated with the SARS-CoV-2 spike (S) protein was used for neutralization assays on Vero cells, as previously described [24]. Briefly, sera were heat-inactivated for 30 minutes at 56°C then brought to an initial 10-fold dilution, followed by four 4-fold dilutions in duplicate. Each dilution was mixed with an equal volume containing 1,250 plaque-forming units of the VSV-S virus and incubated at 37ºC for 1 h. Subsequently, the mixture was added to Vero cells and incubated for 18 hours, after which GFP expression was measured using a live cell microscope system (IncuCyteS3; Sartorius). All sera which did not reduce viral replication by 50% at 1/20 dilution were considered non-neutralizing and were arbitrarily assigned a value of 1/10. All sera that did not result in > 70% recovery of GFP signal at the highest antibody dilution were retested using 5-fold dilutions ranging between 100 and 12,500-fold. Finally, the antibody dilution resulting in 50% virus neutralization (NtAb_50_) was calculated using the drc package (version 3.0-1) in R via a 2 parameter logistic regression model (LL.2 model).

### 3.3. Commercial SARS-CoV-2 IgG immunoassays

Four commercially-available semiquantitative immunoassays were used in the current study. Performance and interpretation of results were done in accordance with the respective manufacturer’s instructions. The LIAISON SARS-CoV-2 S1/S2 IgG chemiluminescent assay (DiaSorin S.p.A., Saluggia, Italy) detects IgG antibodies directed against a recombinant S protein (S1/S2). Samples displaying < 12.0 AU/mL are considered negative, those ranging between 12.0 to 15.0 AU/mL are undetermined and those > 15 AU/mL are deemed as positive. The Euroimmun SARS-CoV-2 IgG ELISA (Euroimmun, Lübeck, Germany) uses a recombinant S1 domain of the S protein as a target. Results are expressed as a ratio, calculated by dividing the optical densities of the sample by those of an internal calibrator provided with the test kit. The cut-off index (COI) for samples to be considered positive was ≥ 1.1 and inconclusive from 0.8 and 1.09. The MAGLUMI 2019-nCoV IgG is an indirect CLIA for assessment of IgG antibodies against SARS-CoV-2 S and nucleocapsid (N) proteins on the fully automated MAGLUMI analyzers (SNIBE – Shenzhen New Industries Biomedical Engineering Co., Ltd, Shenzhen, China). A test result ≥ 1.10 AU/mL is considered positive. The COVID-19 ELISA IgG (Vircell Spain S.L.U., Granada, Spain) is an enzyme immunoassay that detects IgGs targeting the S1 and N proteins. Sera displaying antibody indices (AI) < 1.4 are considered negative, those between 1.4 < 1.6 inconclusive, and those > 1.6 positive. For all the assays, inconclusive results were considered as positives for analysis purposes.

### 3.4. Definitions

Here, NtAb titers ≥1/160 were considered high as this is the minimum NtAb_50_ titer of plasma from COVID-19 convalescent individuals recommended by the FDA for therapeutic use [25]. Trajectories of antibody titers/levels were categorized as ascendant, descendant, fluctuating, or constant. To this end, variations of antibody levels > 10% across sequential specimens were deemed to be relevant, as the intra-run coefficient of variation was below this figure for all the immunoassays (all samples from a given individual were assayed in the same run).

### 3.5. Statistical methods

Test performances were evaluated for their sensitivity with the associated 95% confidence interval (CI). Cohen’s Kappa (κ) statistic was used to evaluate the qualitative agreement between immunoassays. Spearman’s rank test was used to assess the correlation between continuous variables using the entire dataset. To identify the optimal SARS-CoV-2 IgG levels measured by commercial immunoassays predicting NtAb_50_ titers ≥1/160, receiver operating characteristic (ROC) curve analysis was performed. Two-sided exact *P*-values are reported, and a *P*-value < 0.05 is considered statistically significant. The analyses were performed using SPSS version 20.0 (SPSS, Chicago, IL, USA). Kinetics of antibody titers/levels were categorized as ascendant (an increase of antibody levels > 10% compared to previous sampling point), descendant (any decrease), fluctuating, or constant.

## 4. Results

### 4.1. Patients characteristics

All 51 patients presented with pneumonia and imaging or laboratory findings compatible with COVID-19 and were hospitalized in either the pneumology ward (n = 27) or the intensive care unit (ICU; n = 24). As shown in Table 1, most patients (69%) had one or more co-morbidities and displayed high serum levels of several pro-inflammatory biomarkers at the time of serological testing. Four ICU patients eventually died.

### 4.2. Agreement between NtAb and SARS-CoV-2 IgG immunoassays results

Qualitative results returned by immunoassays were evaluated either considering the entire data set or grouping sera according to the time of sampling after the onset of symptoms (< 15 days or ≥ 15 days) (Table 2). As shown in Table 3, overall, results provided by the COVID-19 ELISA IgG test best matched those obtained with the NtAb assay (κ, 0.84; 95% CI, 0.63-1), followed by those of the Euroimmun SARS-CoV-2 IgG ELISA (κ, 0.5; 0.52; 0.52; 95% CI, 0.22-0.81), LIAISON SARS-CoV-2 S1/S2 IgG (κ 95% CI, 0.2-0.78) and MAGLUMI 2019-nCoV IgG (0.4; 95% CI, 0.2-0.77). The same trend was observed when sera collected either < 15 days or ≥ 15 days after the onset of symptoms were analyzed separately. Notably, the concordance between results returned by the NtAb assay and the COVID-19 ELISA IgG was 100% for sera obtained at ≥ 15 days following symptom onset.

**Table 2.** Performance of an antibody neutralization method using a reporter-based pseudotyped virus (vesicular stomatitis virus pseudotyped with the SARSCoV-2 spike protein) and four commercial SARS-CoV-2 IgG immunoassays for the diagnosis of COVID-19

| Qualitative results/Time of sampling after the onset of symptoms<sup>a</sup> | Antibody assay |  |  |  |  |
| --- | --- | --- | --- | --- | --- |
|  | GFP-VSV-SARS-CoV-2 S pseudotype NtAb test | Euroimmun SARS-CoV-2 IgG ELISA | LIAISON SARS-CoV-2 S1/S2 IgG | MAGLUMI 2019-nCoV IgG | COVID-19 ELISA IgG |
| Positive (all sera) | 83 | 76 | 75 | 77 | 83 |
| Negative (all sera) | 7 | 14 | 15 | 13 | 7 |
| Positive/<15 days | 37 | 31 | 30 | 31 | 37 |
| Negative/<15 days | 4 | 10 | 11 | 10 | 4 |
| Positive/≥15 days | 46 | 45 | 45 | 46 | 46 |
| Negative/≥15 days | 3 | 4 | 4 | 3 | 3 |
| <sup>a</sup>A total of 90 sera were included, of which 41 were collected <15 days after the onset of symptoms and 49 afterwards (≥15 days). |  |  |  |  |  |

**Table 3.**
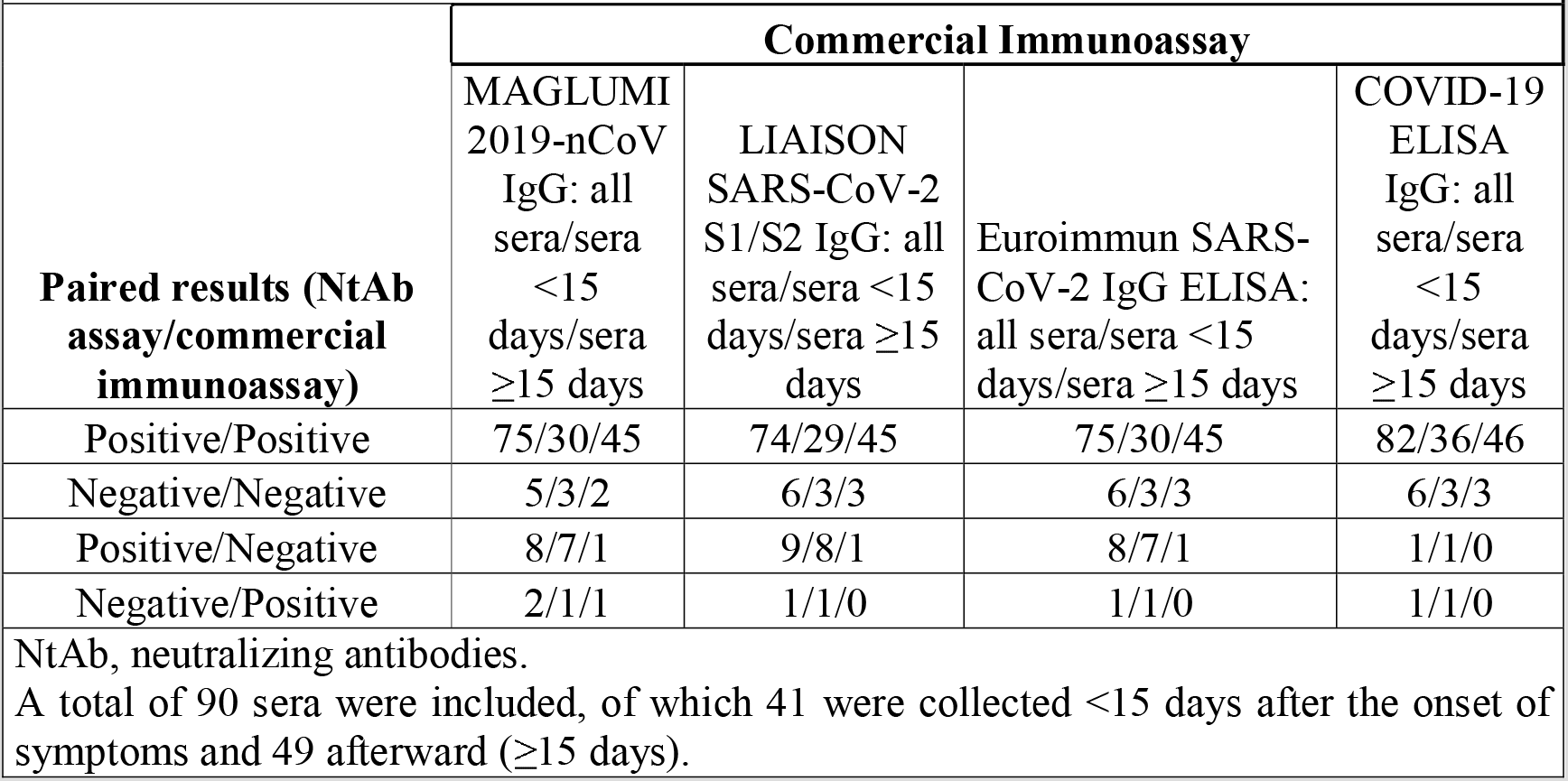
Agreement between the results of a reporter-based pseudotyped virus antibody neutralization method (vesicular stomatitis virus pseudotyped with the SARS-CoV-2 spike protein) and four commercial SARS-CoV-2 IgG immunoassays.

| **Paired results (NtAb assay/commercial immunoassay)** | **Commercial Immunoassay** |  |  |  |
| --- | --- | --- | --- | --- |
|  | MAGLUMI 2019-nCoV IgG: all sera/sera <15 days/sera ≥15 days | LIAISON SARS-CoV-2 S1/S2 IgG: all sera/sera <15 days/sera ≥15 days | Euroimmun SARS-CoV-2 IgG ELISA: all sera/sera <15 days/sera ≥15 days | COVID-19 ELISA IgG: all sera/sera <15 days/sera ≥15 days |
| Positive/Positive | 75/30/45 | 74/29/45 | 75/30/45 | 82/36/46 |
| Negative/Negative | 5/3/2 | 6/3/3 | 6/3/3 | 6/3/3 |
| Positive/Negative | 8/7/1 | 9/8/1 | 8/7/1 | 1/1/0 |
| Negative/Positive | 2/1/1 | 1/1/0 | 1/1/0 | 1/1/0 |

NtAb, neutralizing antibodies.
A total of 90 sera were included, of which 41 were collected <15 days after the onset of symptoms and 49 afterward (≥15 days).

### 4.3. The sensitivity of NtAb and SARS-CoV-2 IgG immunoassays

Overall, the most sensitive tests were the GFP reporter-based pseudotyped virus neutralization assay and the COVID-19 ELISA IgG, followed by the MAGLUMI 2019-nCoV IgG, the Euroimmun SARS-CoV-2 IgG ELISA and the LIAISON SARS-CoV-2 S1/S2 IgG (Table 4). Differences in sensitivity were more apparent when sera were collected early after the onset of symptoms (< 15 days) were analyzed independently, and these tended to decrease in sera obtained at a later time point (Table 4).

**Table 4.**
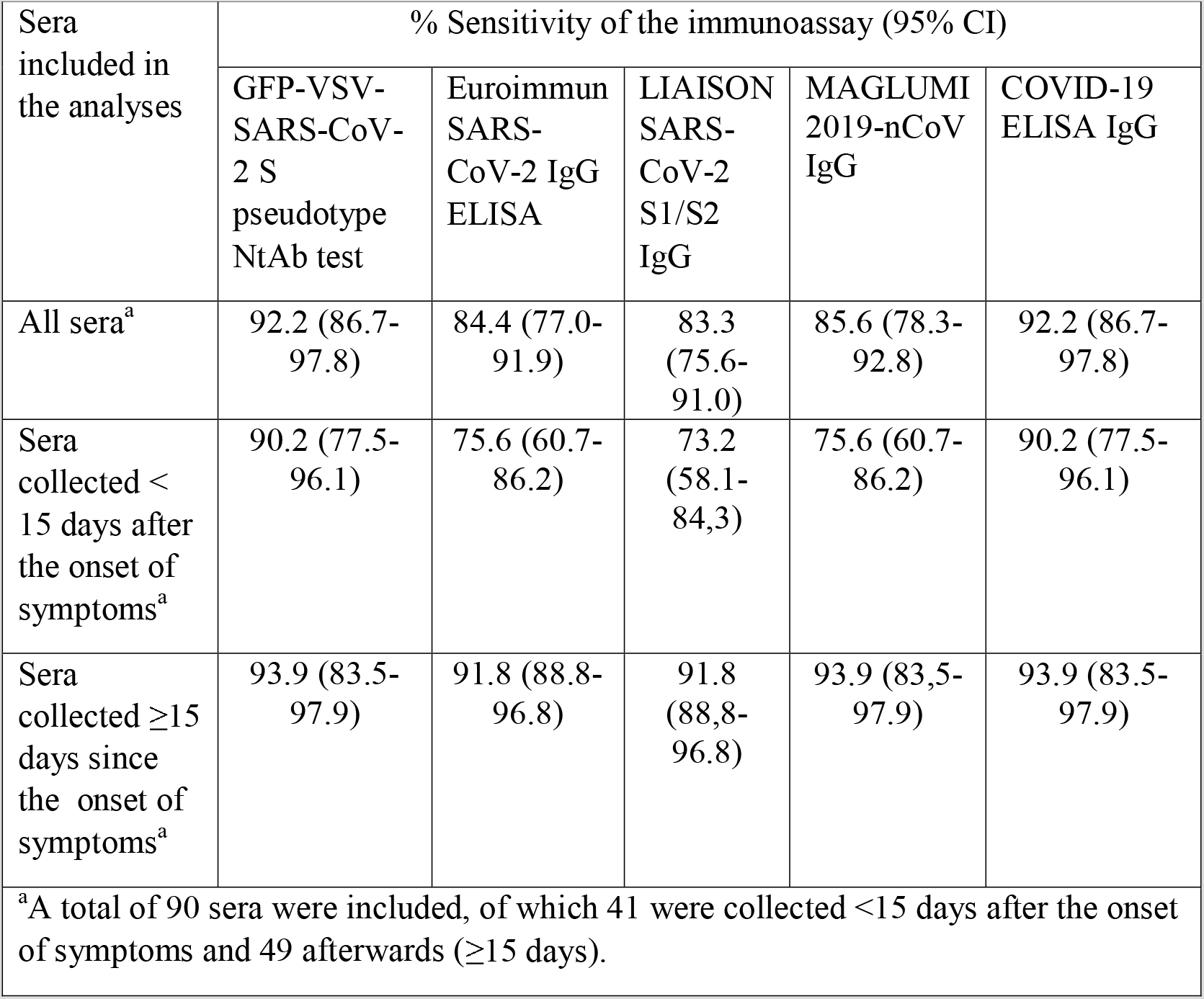
Clinical sensitivity of an antibody neutralization method using a reporter-based pseudotyped virus (vesicular stomatitis virus pseudotyped with the SARS-CoV-2 spike protein) and four commercial SARS-CoV-2 IgG immunoassays for the diagnosis of COVID-19

### 4.4. Correlation between NtAb_50_ titers and IgG levels measured by SARS-CoV-2 IgG immunoassays

NtAb_50_ titers ranged from ≤1/20 (undetectable) to 1/12,500. As shown in Figure 1, overall, the correlation between NtAb_50_ titers and SARS-CoV-2 IgG levels, as returned by the corresponding immunoassay, was strong for the Euroimmun SARS-CoV-2 IgG ELISA (Rho = 0.73) and moderate (Rho = 0.48 to 0.59) for the remaining platforms. In all instances, correlations were positive and statistically significant. The degree of correlation between NtAb_50_ titers and SARS-CoV-2 IgG levels was substantially better for sera collected within the first two weeks after the onset of symptoms (Rho = 0.54-0.80) than for those obtained afterward (Rho = 0.30–0.52), irrespective of the commercial immunoassay employed.

**Figure 1.**
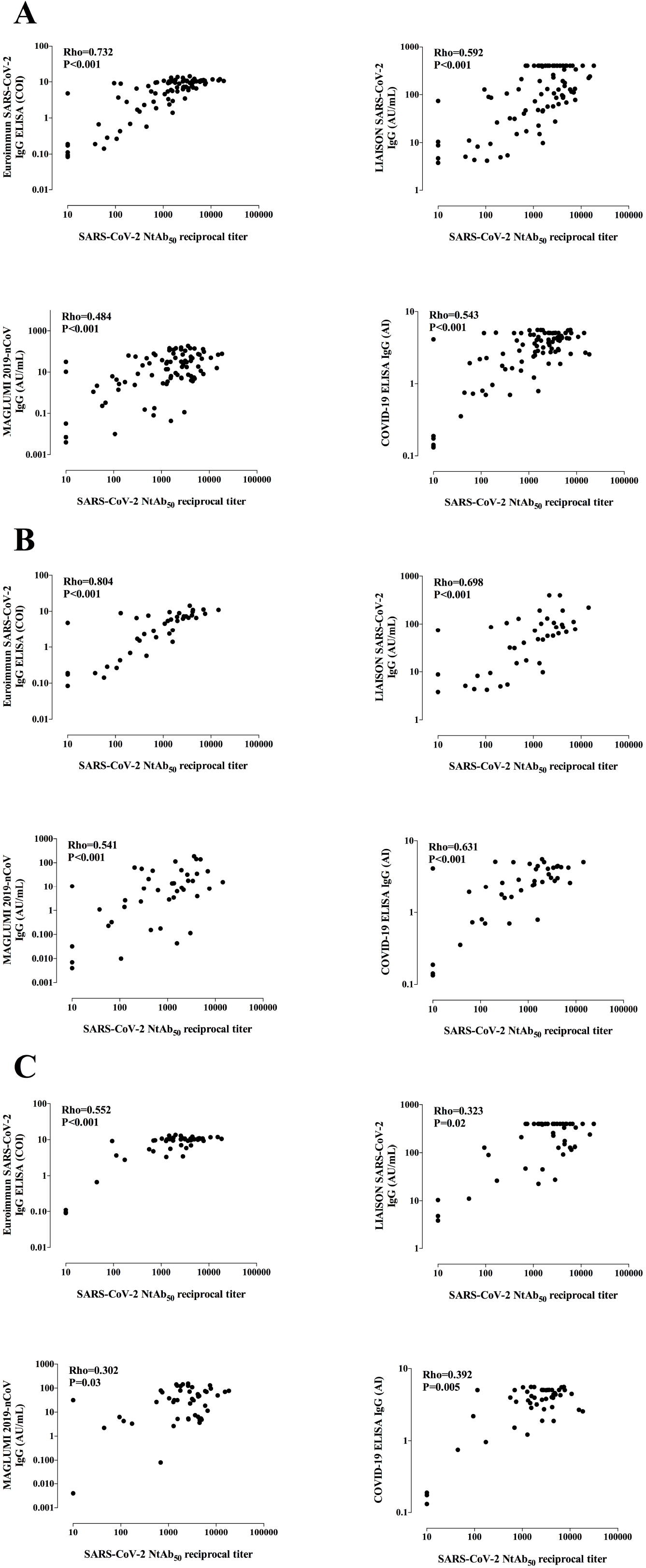
Correlation between NtAb_50_ titers measured by a SARS-CoV-2-S pseudotyped virus neutralization assay and IgG levels measured by commercial SARS-CoV-2 IgG immunoassays in all sera (A), in sera obtained within the first two weeks after the onset of symptoms (< 15 days; B), and in sera collected afterward (≥ 15 days; C). Rho and *P* values are shown.

### 4.5. Inference of high NtAb_50_ titers by using SARS-CoV-2 IgG immunoassays

Seventy-four out of 90 sera displayed high NtAb_50_ titers (≥1/160). ROC analysis was performed to determine the IgG levels measured by commercial immunoassays that predict NtAb_50_ titers of such a magnitude (Figure 2). Specificity was prioritized at the expense of sensitivity for threshold selection. The data are shown in Table 5. The best combination of specificity, sensitivity, and predictive values was achieved by the LIAISON SARS-CoV-2 S1/S2 IgG threshold, with minimal differences across the remaining platforms.

**Figure 2.**
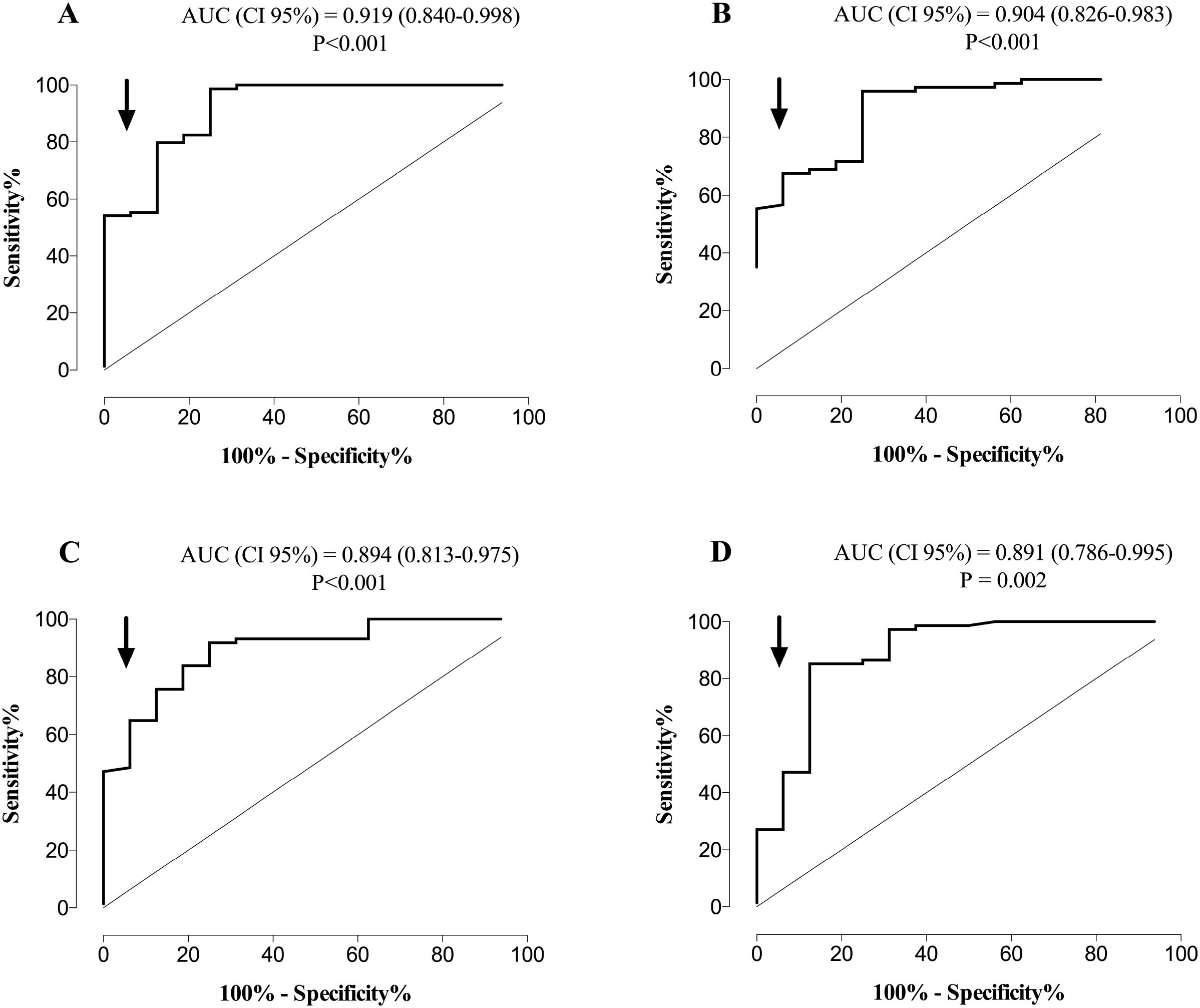
ROC curve analysis for establishing the optimal SARS-CoV-2 IgG threshold levels that predict the presence of high NtAb_50_ titers (≥ 1/160) in hospitalized patients with COVID-19 for the Euroimmun SARS-CoV-2 IgG ELISA (A), LIAISON SARS-CoV-2 S1/S2 IgG assay (B), MAGLUMI 2019-nCoV IgG (C), and COVID-19 ELISA IgG (D) assays.

**Table 5.** SARS-CoV-2-S- IgG threshold levels measured by four commercial immunoassays for predicting neutralizing antibody titers ≥1/160 measured by a reporter-based pseudotyped virus antibody neutralization method (vesicular stomatitis virus pseudotyped with the SARS-CoV-2 spike protein)

| Immunoassay | Threshold | Specificity<br>(CI,95%) | Sensitivity<br>(CI, 95%) | Positive<br>predictive<br>value (CI,<br>95%) | Negative<br>predictive<br>value (CI,<br>95%) |
| --- | --- | --- | --- | --- | --- |
| Euroimmun<br>SARS-CoV-2<br>IgG ELISA | 8.9 COI | 93.8<br>(81.5-100) | 55.4 (44.1-66.7) | 97.6 (87.7-99.6) | 31.2 (19.9-45.3) |
| LIAISON<br>SARS-CoV-2<br>S1/S2 IgG | 90.6 AU | 93.8<br>(81.5-100) | 67.6 (56.9-78.2) | 98.1 (89.7-99.7) | 38.5 (24.9-54.1) |
| MAGLUMI<br>2019-nCoV<br>IgG | 10.9 AU | 93.8<br>(81.5-100) | 64.9 (54.0-75.7) | 98.0 (89.3-99.6) | 36.6 (23.6-51.9) |
| COVID-19<br>ELISA IgG | 4.1 AI | 93.8<br>(81.5-100) | 47.3 (35.9-58.7) | 97.2 (85.8-99.5) | 27.8 (17.6-40.9) |

### 4.6. Longitudinal follow-up of NtAb_50_ titers and SARS-CoV-2 IgG levels measured by immunoassays

We selected patients (n = 11) with 3 or more sera available for kinetic studies. Kinetics of NtAb_50_ titers and SARS-CoV-2 IgG levels determined by commercial immunoassays varied notably across patients and were dependent on the immunoassay considered. Concordance between trajectories of NtAb_50_ titers and SARS-CoV-2 IgG levels was highest when using the COVID-19 ELISA IgG (6 out of 11 patients) assay, followed by MAGLUMI 2019-nCoV IgG (5 out of 11), and LIAISON SARS-CoV-2 S1/S2 IgG and Euroimmun IgG (both 3 out of 11) assays. Representative cases of concordant and discordant trajectories are depicted in Figure 3.

**Figure 3.**
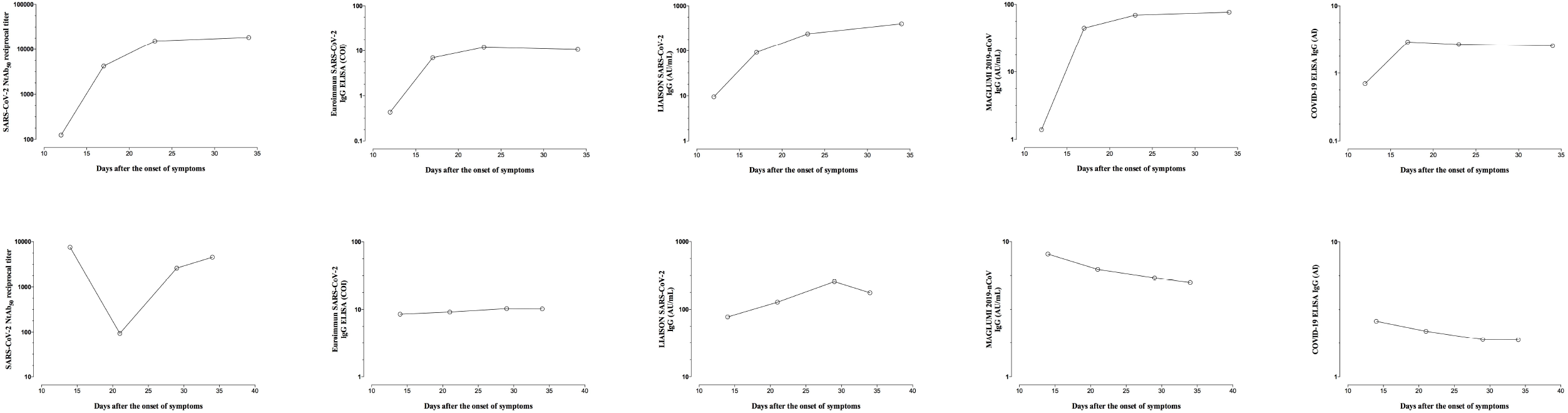
Kinetics patterns of serum NtAb_50_ titers quantitated by a SARS-CoV-2-S pseudotyped virus neutralization assay and SARS-CoV-2 IgG levels measured by commercial immunoassays in two representative patients displaying concordant (upper panels) or discordant (lower panels) kinetics.

## 5. Discussion

A major role in preventing SARS-CoV-2 infection and blunting virus spread in COVID-19 patients is predicted for NtAb [3–5]. Quantitation of serum NtAb titers in plasma from convalescent COVID-19 patients is recommended for optimal selection of specimens for passive transfer therapies; the FDA recommends the use of plasma with NtAb_50_ levels ≥1/160 [22]. NtAb assays, either using SARS-CoV-2 strains, pseudotyped viruses or chimeric viruses are technically demanding, require specialized facilities facilities (biosafety level 3 for live SARS-CoV-2 or level 2 for the pseudotyped viruses), and cannot be implemented in routine practice. Thus, it is of interest to find alternative methods to obtain reliable information regarding the neutralizing activity of sera that are simple, fast, high-throughput assays, and commercially available. Here, we compared the performance of a GFP reporter-based pseudotyped virus neutralization assay (VSV-SARS-CoV-2 S) and four commercial immunoassays targeting the SARS-CoV-2 S protein for the diagnosis of COVID-19 and assessed the extent to which NtAb_50_ titers and SARS-CoV-2 IgG levels measured by these assays correlate. Prior studies assessing this relationship have been published for the Euroimmun SARS-CoV-2 IgG ELISA and LIAISON SARS-CoV-2 S1/S2 IgG [12,13,15,18–20], but not, to the best of our knowledge, for the MAGLUMI 2019-nCoV IgG and COVID-19 ELISA IgG assays. It must be highlighted that antibody titers measured by the NtAb assay used in the current study strongly correlate with those quantitated by assays using wild type SARS-CoV-2 [26].

Here, the overall agreement between qualitative (positive/negative) results returned by the NtAb assay and the four commercially-available immunoassays was fairly good, ranging from 97% for the COVID-19 ELISA IgG assay to 88% for LIAISON SARS-CoV-2 S1/S2 IgG and MAGLUMI 2019-nCoV IgG assays. Concordance across results was better for sera obtained at late times after the onset of symptoms (≥15 days), likely reflecting the existence of differences in the kinetics of NtAb and SARS-CoV-2 IgG levels measured by commercial immunoassays early after infection. In line with this finding, the COVID-19 ELISA IgG performed comparably to the NtAb assay in terms of overall sensitivity (92.2%), ranking first across the evaluated commercial immunoassays. As previously reported, the sensitivity of all immunoassays increased after the second week following the onset of symptoms [12,13,15,16, 18,19, 20]. The overall degree of correlation between NtAb_50_ antibody titers and SARS-CoV-2 IgG levels was strong for the Euroimmun IgG assay but low to moderate for the other commercial immunoassays. The fact that the Euroimmun IgG assay detects antibodies against S1, which contains the RBD domain and concentrates immunodominant epitopes eliciting high-affinity NtAb antibodies [3,5,21,22] may account for this finding. IgG antibodies lacking neutralizing activity and recognizing epitopes within S2 and N proteins (additional targets in the remaining immunoassays) may compete with those binding S1, thus decreasing the degree of correlation. Interestingly, in all cases, correlations were better for sera collected early after the onset of symptoms than for those obtained at later times; in line with this, NtAb trajectory could not be inferred in a large fraction of patients by sequentially monitoring SARS-CoV-2 IgG levels, regardless of the commercial immunoassay used. However, threshold levels of IgGs predicting high NtAb_50_ titers (≥1/160) with high specificity (93%) could be established for all immunoassays, although sensitivities were overall poor.

In the absence of a reference panel of sera with standard levels of NtAb antibodies, comparison across studies addressing the above issue is not straightforward, as the biological characteristics of the neutralization assay, the timing of sera collection and the clinical severity of COVID-19 patients differ notably between them and may explain certain discrepancies. Namely, Weidner et al. [12] reported a strong correlation between IgG levels measured by the LIAISON SARS-CoV-2 S1/S2 IgG and NtAb_50_ titers quantified by a wild type SARS-CoV-2 assay (Rho = 0.75 versus Rho = 0.59 in the current study) in sera from convalescent COVID-19 patients. The degree of correlation reported by Muecksh et al. [20] in a comparable population using an HIV pseudotyped platform was even higher (Rho = 0.82). Likewise, GeurtvanKessel et al. [13] found a strong correlation (Rho = 0.75) using a live SARS-CoV-2 microneutralization assay and sera from a mixed patient population in terms of COVID-19 severity. Despite the above-highlighted differences across studies, our correlation data agree with those of Weidner et al. [12] and GeurtvanKessel et al. [13] for the Euroimmun IgG assay (Rho 0.75 and 0.76, respectively, versus 0.73 herein) and those of Criscuolo et al. [15] who reported a low correlation between IgG levels measured by the LIAISON SARS-CoV-2 S1/S2 IgG and NtAb_50_ titers quantified by using a live SARS-CoV-2 platform in sera obtained within 15 days after the onset of symptoms in a small cohort of COVID-19 hospitalized patients. Moreover, Bonelli et al. [18] found that the probability of having NtAb_50_ titers > 1/160, as determined by a SARS-CoV-2 wild type assay, was 92% when LIAISON SARS-CoV-2 S1/S2 IgG levels were > 80 AU/ml (90 AU/ml in the current study for a positive predictive value of 98%). Also, in line with a previous report [20], the trajectory of NtAb_50_ titers could not be predicted by assessing SARS-CoV-2 IgG levels when measured by commercial immunoassays.

In summary, our data indicate that the reliability SARS-CoV-2 IgG commercial immunoassays targeting the S protein to infer neutralizing activity against this protein in sera from hospitalized COVID-19 patients varies widely across tests, and depends upon time of sera collection after the onset of symptoms.

## Data Availability

Raw data were generated at Hospital Clinico Valencia. Derived data supporting the finding of this study are available from the corresponding author on request

## Ethical Approval

The current study was approved by the Research Ethics Committee of Hospital Clínico Universitario INCLIVA (March 2020).

## Author statement

AV: methodology, analysis of data, validation, review & editing; IT: formal analysis, review and editing; VL: Methodology, investigation; CFG: Methodology, investigation; EA: resources, project administration, review and editing; RG-R: methodology, investigation, validation, funding acquisition, review & editing; MJA: methodology, analysis of data, validation, review & editing; JB: supervision; review & editing; JRD: Conceptualization, supervision, funding acquisition, review& editing; RG: Methodology, investigation, validation, funding acquisition, review & editing; DN: Conceptualization, supervision, writing the original draft, review & editing.

## Acknowledgments

The authors would like to thank Gert Zimmer (Institute of Virology and Immunology, Mittelhäusern/Switzerland), Stefan Pöhlmann and Markus Hoffmann (both German Primate Center, Infection Biology Unit, Goettingen/Germany) for providing the reagents required for the generation of pseudotyped VSV system. Eliseo Albert holds a Río Hortega research contract from the Carlos III Health Institute (Ref. CM18/00221). Ron Geller holds a Ramón y Cajal fellowship from the Spanish Ministry of Economy and Competitiveness (RYC-2015-17517).

## References

1 Zhou F, Yu T, Du R, Fan G, Liu Y, Liu Z, et al Clinical course and risk factors for mortality of adult inpatients with COVID-19 in Wuhan, China: a retrospective cohort study. Lancet 2020; 395: 1054–1062.

2 Guan W-j, Ni Z-y, Hu Y, Liang WH, Ou CQ, He JX, et al Clinical characteristics of coronavirus disease 2019 in China. New Engl J Med 2020; 382: 1708–1720.

3 Moore JP, Klasse PJ. SARS-CoV-2 vaccines: ‘Warp Speed’ needs mind melds not warped minds. J Virol 2020 Jun 26: JVI.01083–20. doi: 10.1128/JVI.01083-20.

4 Zohar T, Alter G. Dissecting antibody-mediated protection against SARS-CoV-2. Nat Rev Immunol 2020; 20: 392–394.

5 Zost SJ, Gilchuk P, Case JB, Binshtein E, Chen RE, Nkolola JP, et al Potently neutralizing and protective human antibodies against SARS-CoV-2. Nature 2020 Jul 15. doi: 10.1038/s41586-020-2548-6

6 Shen C, Wang Z, Zhao F, ang Y, Li J, Yuan J, et al Treatment of 5 critically ill patients with COVID-19 with convalescent plasma. JAMA 2020; 323: 1582–1589.

7 Xia X, Li K, Wu L, Wang Z, Zhu M, Huang B, et al Improved Clinical Symptoms and Mortality on Severe/Critical COVID-19 Patients Utilizing Convalescent Plasma Transfusion. Blood 2020 Jun 23:blood.2020007079.

8 Wang Y, Zhang L, Sang L, Ye F, Ruan S, Zhong B, et al Kinetics of viral load and antibody response in relation to COVID-19 severity. J Clin Invest 2020 Jul 7: 138759.

9 Ni L, Ye F, Cheng M.-L, Deng YQ, Zhao H, Wei P, et al Detection of SARS324 CoV-2-specific humoral and cellular immunity in COVID-19 convalescent individuals. Immunity 2020 doi: 10.1016/j.immuni.2020.04.023.

10 To KK, Tsang OT, Leung WS, am AR, Wu TC, Lung DC, et al Temporal profiles of viral load in posterior oropharyngeal saliva samples and serum antibody responses during infection by SARS-CoV-2: an observational cohort study. Lancet Infect Dis 2020; 20: 565–574.

11 Foundation for Innovative New Diagnostics. SARS-COV-2 Diagnostic Pipeline. https://www.finddx.org/covid-19/pipeline/?section=immunoassays#diag_tab Accessed July 20, 2020.

12 Weidner L, Gänsdorfer S, Unterweger S, Weseslindtner L, Drexler C, Farcet M, et al Quantification of SARS-CoV-2 antibodies with eight commercially available immunoassays. J Clin Virol 2020 Jul 6; 129: 104540. doi: 10.1016/j.jcv.2020.104540.

13 GeurtsvanKessel CH, Okba NMA, Igloi Z, Bogers S, Embregts CWE, Laksono BM, et al An evaluation of COVID-19 serological assays informs future diagnostics and exposure assessment. Nat Commun 2020; 11: 3436.

14 Ng D, Goldgof G, Shy B, Levine A, Balcerek J, Bapat SP, et al SARS-CoV-2 seroprevalence and neutralizing activity in donor and patient blood from theSan Francisco Bay Area. medRxiv preprint 2020. doi:https://doi.org/10.1101/2020.05.19.20107482.

15 Criscuolo E, Diotti RA, Strollo M, Rolla S, Ambrosi A, Locatelli M, et al Poor correlation between antibody titers and neutralizing activity in sera from SARS-CoV-2 infected subjects. medRxiv 2020; doi: https://doi.org/10.1101/2020.07.10.20150375.

16 Jääskeläinen AJ, Kuivanen S, Kekäläinen E, Ahava MJ, Loginov R, Kallio-Kokko H, et al Performance of six SARS-CoV-2 immunoassays in comparison with microneutralisation. J Clin Virol 2020; 129: 104512. doi: 10.1016/j.jcv.2020.104512.

17 Suhandynata RT, Hoffman MA, Huang D, Tran JT, Kelner MJ, Reed SL, et al Commercial Serology Assays Predict Neutralization Activity Against SARS-CoV-2.medRxiv 2020. doi: https://doi.org/10.1101/2020.07.10.20150946.

18 Bonelli F, Sarasini A, Zierold C, Calleri M, Bonetti A, Vismara C, et al Clinical And Analytical Performance Of An Automated Serological Test That Identifies S1/S2 Neutralizing IgG In COVID-19 Patients Semiquantitatively. J Clin Microbiol. 2020 Jun 24:JCM.01224–20. doi: 10.1128/JCM.01224-20

19 Kohmer N, Westhaus S, Rühl C, Ciesek S, Rabenau HF. Brief clinical evaluation of six high-throughput SARS-CoV-2 IgG antibody assays. J Clin Virol. 2020 Aug;129: 104480. doi: 10.1016/j.jcv.2020.104480.

20 Muecksch F, Wise H, Batchelor B, Squires M, Semple E, Richardson C, et al Longitudinal analysis of clinical serology assay performance and neutralising antibody levels in COVID19 convalescents. medRxiv. 2020 Aug 6:2020.08.05.20169128. doi: 10.1101/2020.08.05.20169128

21 Barnes CO, West AP Jr, Huey-Tubman KE, Hoffmann MAG, Sharaf NG, Hoffman PR, et al Structures of Human Antibodies Bound to SARS-CoV-2 Spike Reveal Common Epitopes and Recurrent Features of Antibodies. Cell 2020; S0092-8674(20)30757-1.

22 Rogers TF, Zhao F, Huang D, Beutler N, Burns A, He WT, et al Isolation of Potent SARS-CoV-2 Neutralizing Antibodies and Protection From Disease in a Small Animal Model Science. science 2020; eabc7520. doi: 10.1126/science.abc7520.

23 Giménez E, Albert E, Torres I, Remigia MJ, Alcaraz MJ, Galindo MJ, et al SARS-CoV-2-reactive interferon-γ-producing CD8+ T cells in patients hospitalized with coronavirus disease 2019. J Med Virol 2020 Jun 24. doi: 10.1002/jmv.26213.

24 Gozalbo-Rovira R, Gimenez E, Latorre V, Frances-Gomez C, Albert E, Buesa J, et al SARS-CoV-2 antibodies, serum inflammatory biomarkers and clinical severity of hospitalized COVID-19 Patients. medRxiv 2020. 07.22.20159673; doi: https://doi.org/10.1101/2020.07.22.20159673.

25 Recommendations for Investigational COVID-19 Convalescent Plasma. https://www.fda.gov/vaccines-blood-biologics/investigational-new-drug-ind-or384device-exemption-ide-process-cber/recommendations-investigational-covid-19-convalescent-plasma. Accesed July 5, 2020.

26 Schmidt F, Weisblum Y, Muecksch F, Hoffmann HH, Michailidis E, Lorenzi JCC, et al Measuring SARS-CoV-2 neutralizing antibody activity using pseudotyped and chimeric viruses. J Exp Med. 2020 Nov 2;217(11): e20201181.

